# Generation of Inhibitory Autoantibodies to ADAMTS13 in Coronavirus Disease 2019

**DOI:** 10.1101/2021.03.18.21253869

**Authors:** Adrian A. N. Doevelaar, Martin Bachmann, Bodo Hölzer, Felix S. Seibert, Benjamin J. Rohn, Oliver Witzke, Ulf Dittmer, Thorsten Brenner, Krystallenia Paniskaki, Serap Yilmaz, Rita Dittmer, Sonja Schneppenheim, Nina Babel, Ulrich Budde, Timm H. Westhoff

## Abstract

**Objectives:** It has recently been shown that von Willebrand factor (vWf) multimers may be a key driver of immunothrombosis in Coronavirus disease 2019 (COVID-19). Since COVID-19 is associated with an increased risk of autoreactivity, the present study investigates, whether the generation of autoantibodies to ADAMTS13 contributes to this finding.

**Design:** Observational prospective controlled multicenter study.

**Setting:** Blood samples and clinical data of patients with COVID-19 were collected regularly during hospitalization in the period from April to November 2020.

**Patients:** 90 patients with confirmed COVID-19 of mild to critical severity and 30 healthy controls participated in this study.

**Measuerements and Main Results:** Antibodies to ADAMTS13 occurred in 31 (34.4%) patients with COVID-19. Generation of ADAMTS13 antibodies was associated with a significantly lower ADAMTS13 activity (56.5%, interquartile range (IQR) 21.25 vs. 71.5%, IQR 24.25, p=0.0041), increased disease severity (severe or critical disease in 90% vs. 62.3%, p=0.0189), and a trend to a higher mortality (35.5% vs. 18.6%, p=0.0773). Median time to antibody development was 11 days after first positive SARS-CoV-2-PCR specimen.

**Conclusion:** The present study demonstrates for the first time, that generation of antibodies to ADAMTS13 is a frequent finding in COVID-19. Generation of these antibodies is associated with a lower ADAMTS13 activity and an increased risk of an adverse course of the disease suggesting an inhibitory effect on the protease. These findings provide a rationale to include ADAMTS13 antibodies in the diagnostic workup of SARS-CoV-2 infections.

## Brief report

Coronavirus disease 2019 (COVID-19) is associated with micro- and macrovascular thrombotic events - a phenomenon, which has recently been described as “immunothrombosis”. Macrovascular events comprise both venous thrombembolism and arterial thrombotic events including myocardial infarction, stroke, and limb ischemia. [1] Microvascular thrombosis has preferentially been described by autopsy studies in the lungs.[2]

We and others observed thrombotic microangiopathy (TMA) in a subset of patients.[3] Recently, we demonstrated that COVID-19 is associated with a substantial increase in Von Willebrand Factor (vWf) concentrations, which can exceed the ADAMTS13 processing capacity resulting in the formation of large vWf multimers identical to thrombotic thrombocytopenic purpura (TTP).[4] The ADAMTS13/vWf Antigen (vWf:Ag) ratio was thereby an independent predictor of severity of disease and mortality. In the present study we investigated, whether the generation of antibodies to ADAMTS13 might contribute to this observation.

We performed an observational prospective controlled multicenter study and enrolled 120 participants including 90 patients, who were hospitalized for COVID-19. Patients were recruited at Ruhr-University Bochum, University of Duisburg-Essen, and Asklepios Klinikum Hamburg Harburg, Germany. The severity of COVID-19 ranged from mild to critical and was categorized according to the guidelines of the Robert Koch Institute, Germany. Thirty healthy subjects served as control. The study was approved by the ethical committees of Ruhr-University Bochum (20-6886), University Hospital Essen (20-9214-BO) and the Medical Association Hamburg. Demographic, clinical and hemostaseologic characteristics of patients at initial sample obtainment are summarized in Table 1. ADAMTS13 activity and antibodies to ADAMTS13 were analyzed from citrate-plasma and serum using Technozym ADAMTS13 ELISA (Technoclone, Vienna, Austria). An ADAMTS13 antibody concentration of ≥16 U/mL was considered positive. vWf:Ag was measured using a sandwich ELISA with polyclonal antibodies. Parts of the study population and the control group have previously been described. [4]

**Table 1:**
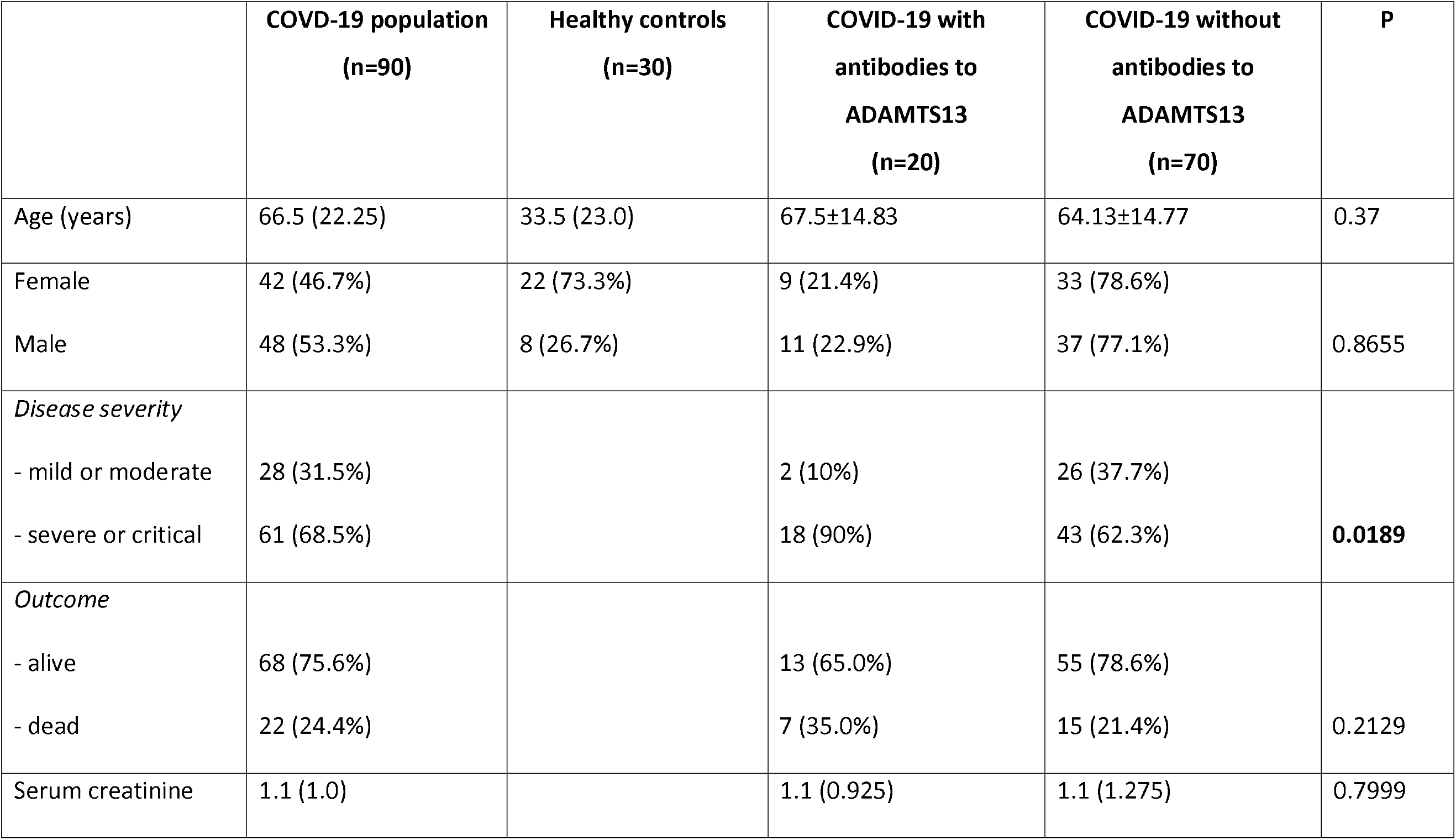
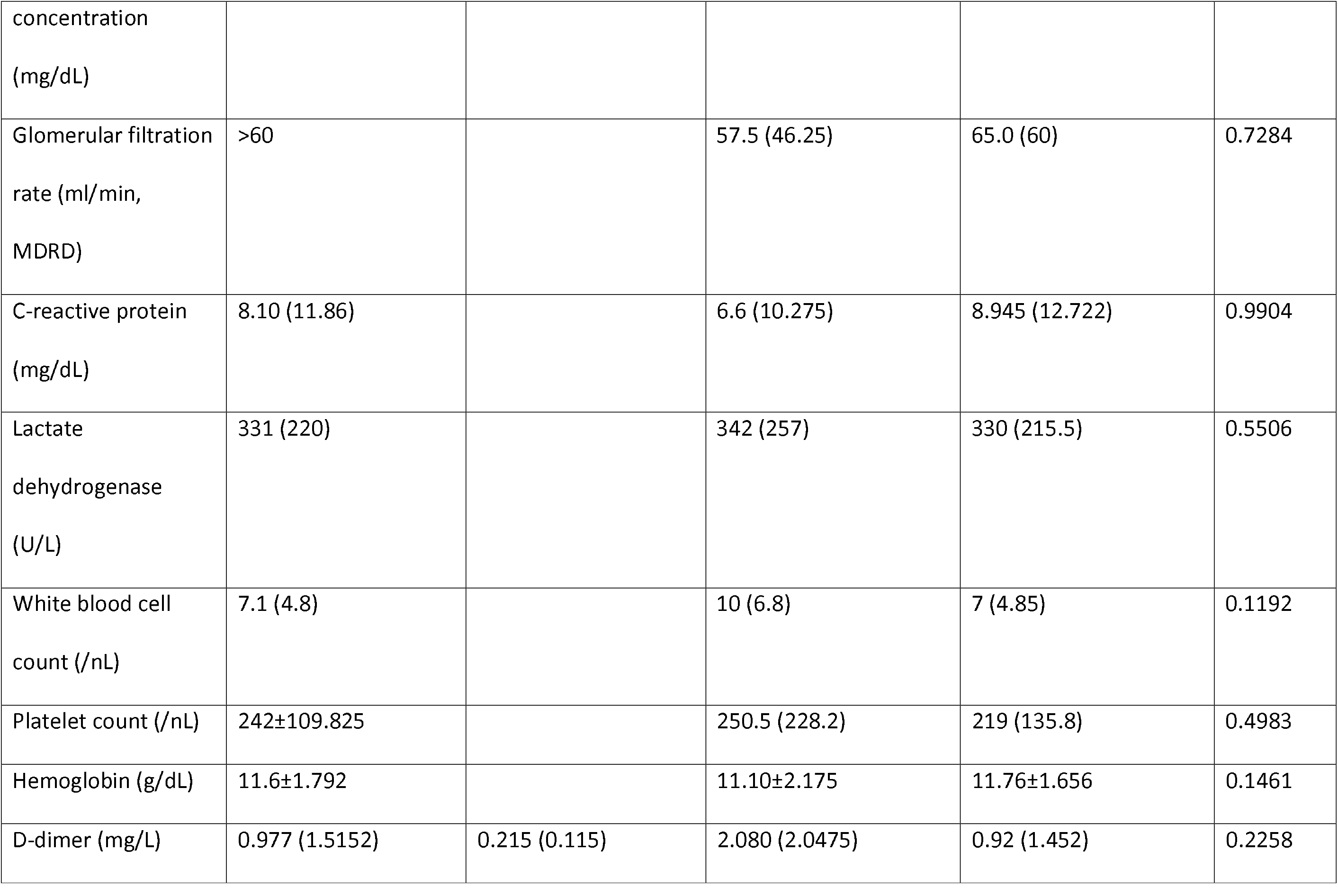
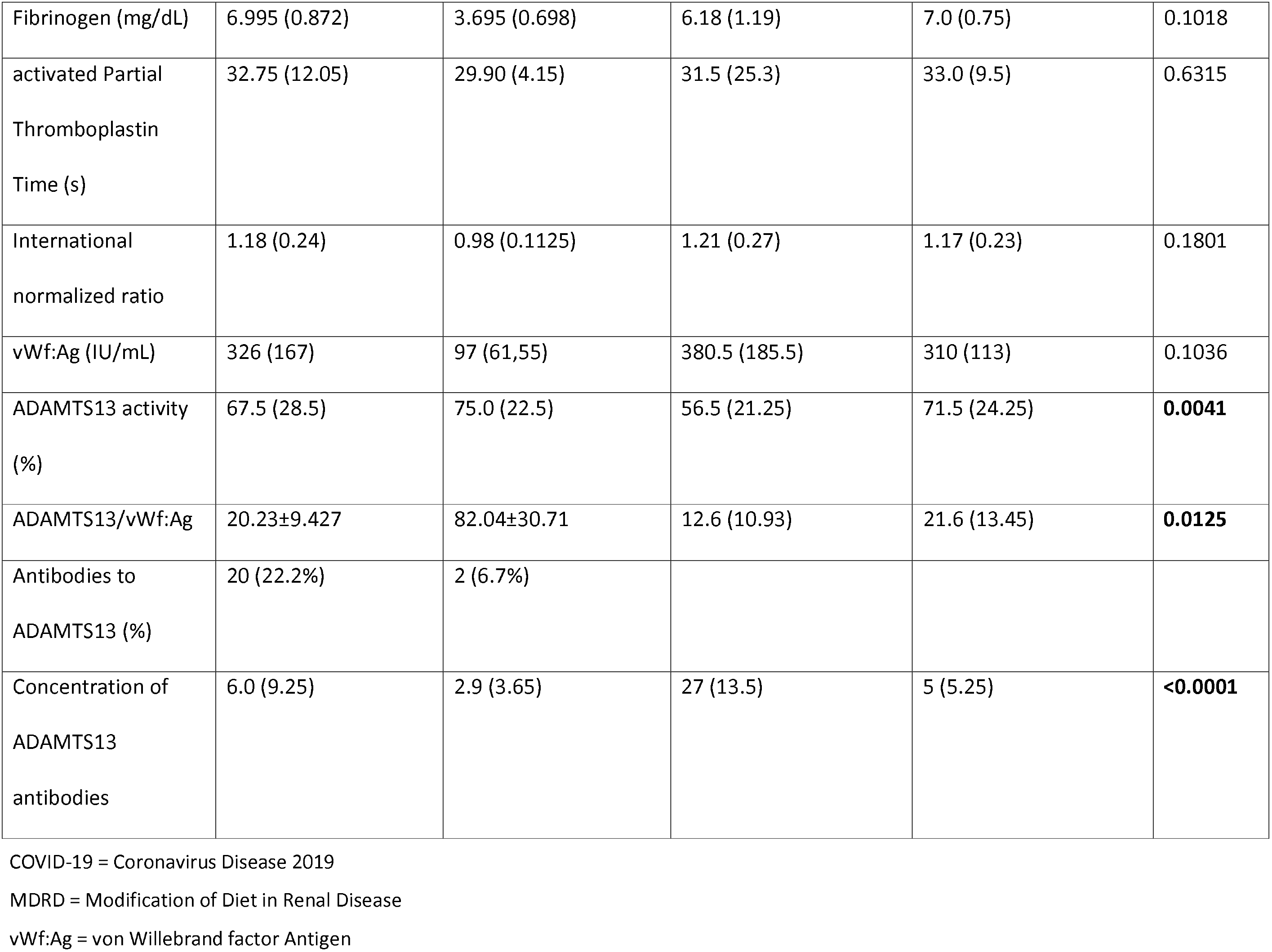
Baseline epidemiological, clinical and hemostaseological characterization of the study population. Data are presented as mean ± standard deviation for normally distributed parameters, otherwise in median and interquartile range. P<0.05 was regarded significant (bold type).

Median time to initial sample obtainment was 4 days after first positive PCR, and 3 days after admission, respectively. VWF:Ag (iU/mL) was significantly higher in patients with COVID-19 than in healthy controls (326 iU/mL, interquartile range (IQR) 163 vs. 97 iU/mL, IQR 61, p<0.0001). Median ADAMTS13 activity was 67.5%, IQR 28.5, in COVID-19 patients and 75.5%, IQR 22.5, in the control group (p=0.0168). The ADAMTS13/VWF:Ag ratio was substantially lower in COVID-19 than in the control group (20.2±9.4 vs. 82.0±30.7, p<0.0001). Twenty patients with COVID-19 (22.2%) had antibodies to ADAMTS13 at initial sample obtainment compared to 2 patients in the control group (6,7%, chi squared p=0.0565). Median concentration of ADAMTS13 antibodies was 27 U/mL (IQR 13.5). In 37 (41.1%) patients, follow up blood samples were available (3 in median, IQR 2). Of these patients, 4 (10.8%) had antibodies to ADAMTS13 at initial obtainment and 11 (29.7%) patients converted from negative to positive. Overall, 31 patients (34.4%) had antibodies at baseline evaluation or developed such during the course of hospitalization (figure 2D).

**Figure 2:**
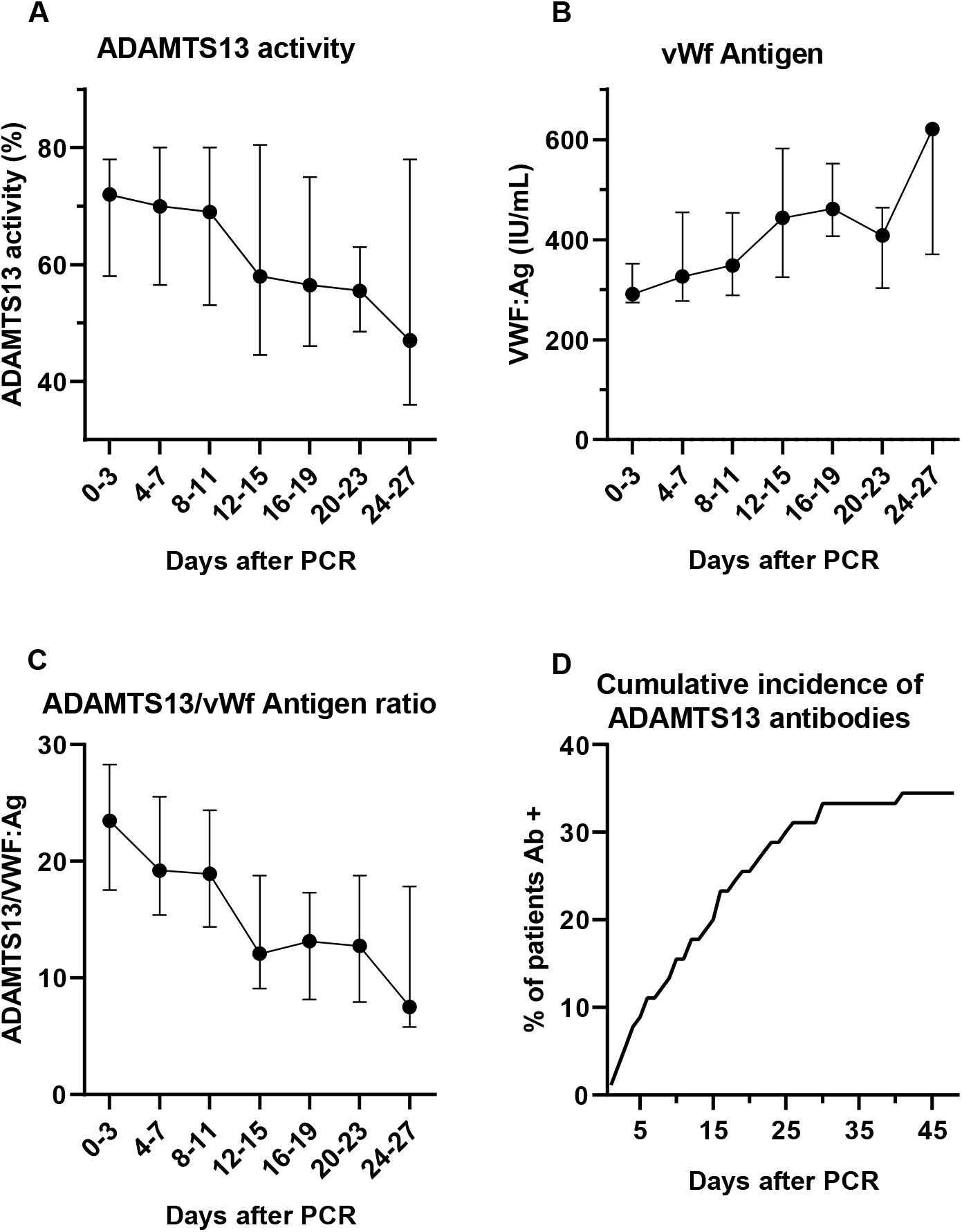
(A) ADAMTS13 activity, (B) von Willebrand factor Antigen (vWf:Ag), (C) ADAMTS13/vWf:Ag ratio and (D) cumulative incidence of ADAMTS13 antibodies (Ab) in the course of Coronavirus disease. Data are presented in median and interquartile range.

Median time to detection of antibodies was 11 days after first positive PCR and 9 days after admission, respectively. At initial sample obtainment median ADAMTS13 activity was significantly lower in those patients with ADAMTS13 antibodies compared to those patients without antibodies (56.5%, IQR 21.25 vs. 71.5%, IQR 24.25, p=0.0041, Figure 1A). Hematological parameters did not significantly differ in presence or absence of ADAMTS13 antibodies.

**Figure 1:**
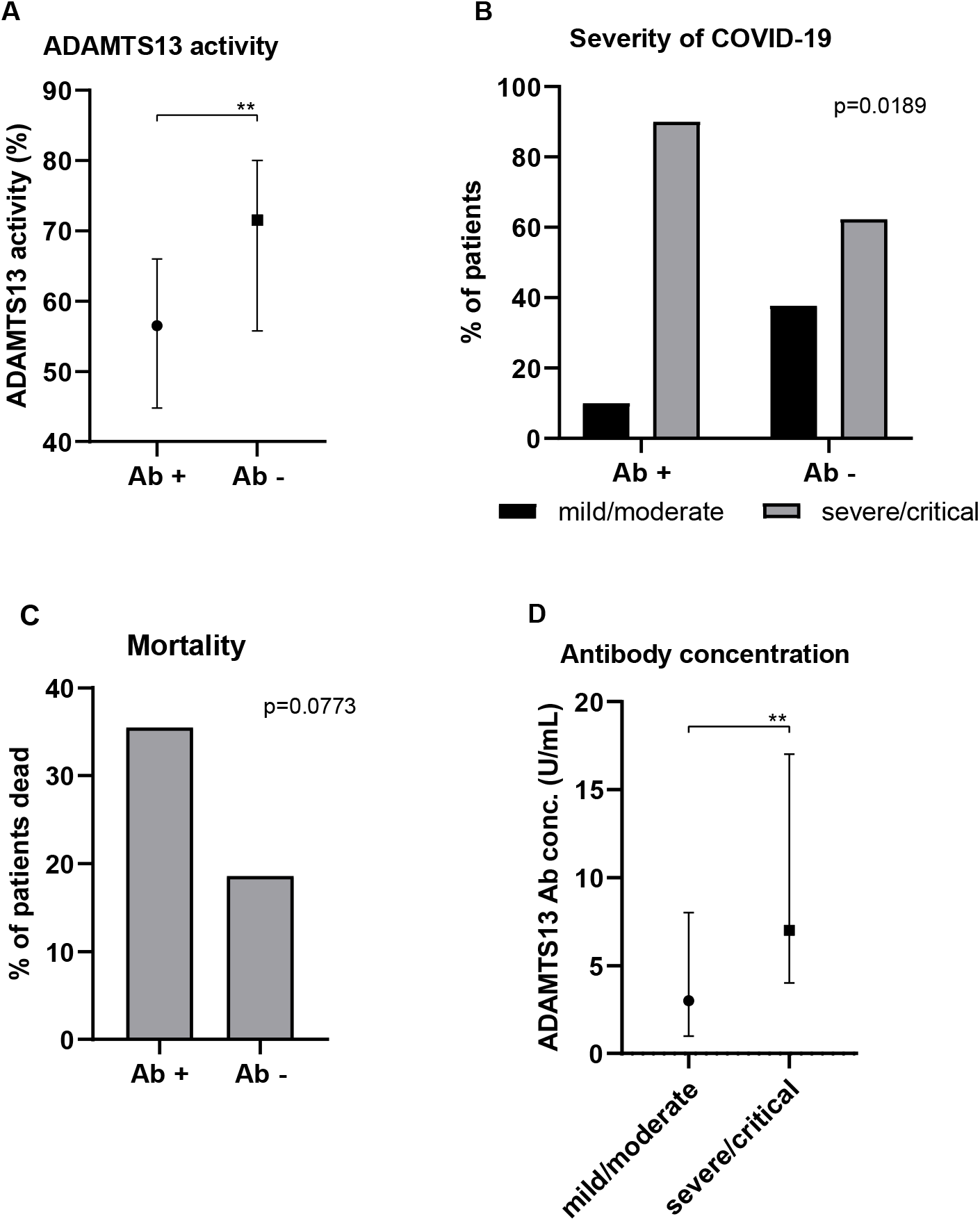
(A) ADAMTS13 activity (%), (B) severity of disease and (C) mortality in dependence on antibody (Ab) status. (D) Ab concentration (conc., U/mL) in dependence on disease severity. Data are presented in % of patients with chi squared p-values, or median and interquartile range, respectively.

Severity of disease differed in dependence of antibody development. Patients who had antibodies at baseline evaluation had a significantly more severe course of disease compared to subjects without antibodies (chi squared p=0.0189, Figure 1B). Patients who developed antibodies against ADAMTS13 during hospitalization tended to have a worse outcome (35.5% vs. 18.6%, chi squared p=0.0773, Figure 1C). Moreover, baseline ADAMTS13 antibody concentration differed significantly among the individual categories of disease severity (Figure 1D). In univariate binary logistic regression analyses baseline ADAMTS13 antibody concentration (regression coefficient (r)=-0.04, p=0.046), ADAMTS13 activity (r=0.038, p=0.009), vWf:Ag (r=-0.006, p=0.001) and ADAMTS13/vWf:Ag ratio (r=0.114, p=0.001) had a significant impact on mortality. In follow-up samples median ADAMTS13 activity and ADAMTS13/vWf:Ag ratio decreased over time (Figure 2A, C), with increasing vWf:Ag levels (figure 2B).

In summary, the present findings show that 1) the higher vWf concentrations and the lower ADAMTS13, the higher the probability of a severe course of COVID-19 including risk of death. 2) The present study demonstrates for the first time, that generation of antibodies against ADAMTS13 is a frequent finding in COVID-19 occurring in approximately one third of hospitalized patients and is associated with a lower ADAMTS13 activity suggesting an inhibitory effect on the protease. 3) The study shows that not only the presence but also the concentration of ADAMTS13 antibodies predicts the severity of COVID-19.

Lower ADAMTS13 activity and ADAMTS13/vWf:Ag ratio have recently been associated with an increase in morbidity and mortality.[4] This finding may in part be attributable to development of these inhibitory antibodies. In general, an ADAMTS13 antibody concentration of ≥16 U/mL is considered positive and diagnostic for TTP. Noteworthy, none of the patients developed severe thrombopenia < 50.000/µl. Thus, the inhibitory effect is likely to be weaker than in TTP.

The immune response to SARS-CoV-2 is associated with an increased risk of autoreactivity. Hence, antibodies to phospholipids and interferon have been described during the pandemic.[5, 6] Recently, lupus- and rheumatoid arthritis-like antibody patterns have been observed in COVID-19.[7] Accordingly, critically ill patients with COVID-19 display hallmarks of extrafollicular B cell activation and shared B cell repertoire features typical of autoimmune settings.[8] Interestingly, the proportion of patients developing an ANA titer of ≥1:160 was very similar (35,6%) to the proportion of subjects developing ADAMTS13 antibodies in the present study (34.4%). SARS-CoV-2 thereby increases the risk of thrombotic microangiopathy by two synergistic mechanisms: First, the ubiquitous endothelial damage induced an excessive release of vWf, which might even exceed the protease activity of physiological concentrations of ADAMTS13. Second, the SARS-CoV-2 induced autoreactive inflammatory milieu leads to the generation of autoantibodies to ADAMTS13, which reduces ADAMTS13 activity and thereby further impairs the protease’s capacity. Both of these mechanisms yield an increased risk of intravascular large and ultralarge vWf multimers with thrombotic microangiopathy resembling TTP.

What are the therapeutic consequences of this study? Plasma exchange constitutes a therapeutic option, which would be able to reduce both the excessive vWf and the antibodies to ADAMTS13 and – moreover – would deliver ADAMTS13. It could thereby reestablish the physiological balance between vWf and its protease. In first case series plasma exchange was used to attenuate circulating cytokines and inflammatory mediators in critically ill patients with COVID-19. Case series from Barcelona and Heidelberg describe favorable effects on parameters of inflammation and clinical outcome[9, 10]. In a cohort in Oman, 11 critically ill patients underwent plasma exchange. Plasma exchange was associated with higher extubation rates and lower mortality.[11] Our findings provide a rationale beyond the elimination of cytokines to suggest plasma exchange as a promising therapeutic strategy in COVID-19.

In conclusion, the present study shows that a substantial part of patients with COVID-19 develop autoantibodies to ADAMTS13 in the course of their disease. Occurrence of antibodies is associated with a significantly lower ADAMTS13 activity, which causes a decreased degeneration of large and ultralarge vWf multimers likely contributing to SARS-CoV-2 induced immunothrombosis. The impact of each individual parameter on morbidity and mortality of COVID-19 is consistent with the physiology of this primary hemostasis system and very much in line with the pathophysiology of thrombotic microangiopathy: The excess of vWf, the decrease of ADAMTS13, the occurrence and the concentration of antibodies to ADAMTS13: Each of these individual parameters independently predicts adverse outcome. The findings that the antibodies predict outcome in a dose-dependent manner and that they are associated with impaired ADAMTS13 activity strongly suggest that these antibodies are indeed of pathophysiological relevance. These findings provide a rationale to consider plasma exchange as a therapeutic option in COVID-19 and to include vWf, ADAMTS13 activity, and antibodies to ADAMTS13 in the diagnostic workup.

## Data Availability

No referral to additional Data or supplementary material.

## Acknowledgements

We thank the laboratory staff Kerstin Will, Claudia Fiedelschuster, Barbara Schocke and Dr. Antje Pieconka of MEDILYS, Hamburg, for their indefatigable efforts in this study in times of excessive routine workloads.

